## supplemental material for "Endemic and epidemic human alphavirus infections in Eastern Panama; An Analysis of Population-based Cross-Sectional Surveys"

### **Additional details on the 2017 sero-survey**

#### *Mogue*

The community of Mogue, on the banks of the Mogue River (Figure 1) in the Province of Darien, which is the home of the Parque Nacional del Darien, the largest national park in Central America and a UNESCO Biosphere Reserve. Yearly rainfall ranges between 1,800 and 4,500mm. The mean maximum temperature is 28°C during the rainy season and 31°C in the dry season 20. The main subsistence activities are fishing, corn cultivation, and tourism. The total population of Mogue is estimated to be around 300 inhabitants.

***Details about recruitment***

In most instances, nonparticipation was due to the inability of researchers to locate the household residents. Residents 1 year of age or older were eligible for inclusion. Each participant was interviewed using a standardized epidemiological form to record occupation, activities, livestock and crop holdings. Older adults and children were interviewed with the help of a close family member when needed. Household-level information (e.g. house structure) was observed and recorded directly by the interviewer where possible. Trained phlebotomists collected 10 ml of blood from persons 9 years of age and older (3 ml for children 1–8 years-old) by peripheral venipuncture using standard aseptic technique. The samples were processed on-site within 6 hours by centrifugation to separate serum, then stored in liquid nitrogen and transported to the GMI for laboratory testing.

***Details about risk factors questionnaire***

Independent variables included age, sex, main occupation, activities including breeding poultry, fishing, cutting bushes, walking/playing in paddocks or crops, working in agriculture, paddocks, grain deposits, sawmills/forest, chicken coops, and pigsties, washing in rivers and bathing. Self-reported symptoms were recorded and included weakness, fatigue, difficulty focusing, memory loss, confusion, dizziness, seizures, fatigue, paralysis, difficult walking, headache, insomnia, depression, irritability, difficulty cooking, difficulty cleaning, difficulty working, fever, chills, vomit, and diarrhea. Other variables related to the house features were floor, wall, roof and window materials, types of crops grown, shrubs surrounding the home, waste management, and water supply.

### **Additional details on the 2012 sero-survey**

For the 2012 survey sites were selected accordingly with previous report of human or equine encephalitis cases due to VEEV of MADV. This survey included the following areas seven areas:

#### *Pijibasal*

The town of Pijibasal is located within the district of Pinogana, 12 km South East of El Real de Santa María. It is surrounded by fields and the Pirre River on the western margin. This town has a grid arrangement, delimited by a concrete path that connects the houses and the school in the periphery. The houses are elevated two meters above the ground, using the ground below them as a deposit for storage chicken coops. A total of 63 inhabitants of the Embera ethnic group make up this town, which is linked by means of a dirt road, the landscape is made up of paddocks and stubble on both banks, it is divided by 4 streams (only one bridge of cement is available in the first) without prominent elevations in the field. The ecosystem between El Real and Pijivasal is similar along the road.

#### *Pirre 2*

Located on the road that connects El Real and Pijivasal. The village is composed of 21 inhabitants of countryside origin, dedicated to agriculture and livestock. Their homes are built with wood, palms and zinc. Some people work cutting wood in the forest and in paddocks with native and improved pastures, plus rice, corn, yucca and plantain crops for family subsistence.

#### *Pirre 1*

On the side of the road is this town with 56 inhabitants of countryside origin. Like Pirre 2, paddocks and stubble are observed on both banks. In some houses, they raise pigs and chickens in small pens, as well as large paddocks in the periphery, although with a small amount of livestock.

#### *Mercadeo*

Mecadeo is located on the banks of the Tuira River, its population is 132 inhabitants of the Embera ethnic group, its houses are grouped along a path and built with wood and stalks, raised on the ground about two meters. In the surroundings of the town we find stubble and subsistence crops of rice, corn, plantain and yucca.

#### *El Real*

The inhabitants are of Negroid, Embera and Latin origin. We can find houses built with different styles and materials (wood, blocks, zinc, etc.). Its population according to the 2010 census is made up of 555 inhabitants. They work at fishing, trade and other informal activities. Access is by boat through the Chucunaque River, the Tuira River and the Pirre River. The surroundings of the town are made up of large areas of paddocks, composed of native grasses and improved for the breeding of cattle. In these grazing areas there are scattered timber and fruit trees. On the edges of the rivers and streams the gallery forests have native trees that do not exceed 30 meters in height.

#### *Tamarindo*

This community is located about 12 km from the village of Santa Fe, District of Chepigana, province of Darién. Most of its inhabitants come from the central provinces specifically Herrera and the South of Veraguas. There are 203 inhabitants according to the 2012 census. Many are engaged in agriculture as the main source of income; the livestock area is minimal.

There are paddocks with native and improved grass, maize and subsistence rice crops, as well as small plots of yucca and fruit trees. There is also some scattered stubble with trees that do not exceed 15 meters in height. There are also small patches of forest on the slopes of the hills that surround the town and gallery forest in the rivers and streams.

#### *Aruza*

Aruza is a community located in the township of Rio Iglesias, district Chepigana close to Metetí and located within three protected areas by law 1) Filo del Tallo Hydrological Reserve, 2) Canglón Forest Reserve and 3) Matusagaratí Lagoon. The population is about 154 inhabitants. The Aruza area is surrounded by paddocks with native grass in the flat areas, corn crops and subsistence rice in the garden of houses, in addition, plots less than one hectare of yucca and banana. There is a small amount of stubble with trees that do not exceed 15 meters in height. Part of the Laguna de Matusagaratí wetland has been invaded and converted into paddocks and rice and oil palm cultivation areas. The secondary forests surrounding the town are located in the highlands and the lowlands are flooded in the rainy season.

### **Additional details on the Force of Infection (FOI) Analysis**

Models were fitted on a Bayesian framework using Stan’s No-U-Turn Sampler [1] with four Markov chains and 20,000 iterations on each and with 50% of these iterations discarded as “warm-up”.

#### *Prior distributions*

***Prior distribution for constant FOI model***

The prior distribution for the FOI estimate in the constant model is based on a uniform distribution

$$FOI \sim uniform(0,2)$$

***Prior distributions for time-varying FOI model***

The prior distribution for FOI estimate in the time-varying FOI model follows a $student t$distribution informed by the FOI estimate from the previous decade:

$$FOI \sim student t (\nu,FOI_{previous decade}, \sigma)$$

$$\sigma\sim Cauchy (0, 1)$$

$$\nu\sim Cauchy (0, 1)$$

$$FOI_{first decade} \sim normal \left( 0, 1 \right)$$

#### *Convergence and Posterior Predictive Checks*

Convergence was assessed by the use of $Rˆ$statistic, which measures the “within chain” variability (W) and compares to the “between chains” variability (B):

$$Rˆ =\sqrt{\frac{W + \frac{1}{n} (B - W ) W}{W}}$$

This method assumes that the chains have been simulated in parallel, each with different starting points, which are overdispersed with respect to the target distribution.

If this metric is large, this suggests that either estimate of the variance can be further decreased by more simulations[2].

It is expected that in convergence, B → W and thus Rˆ → 1.

A value of $Rˆ <1.1$ was considered enough to achieve convergence. We show the convergence plots for the two instances where a time-varying FOI model fit the data the best (See Figures S2 and S3).

We also performed posterior predictive checks and examined the residuals (See Figures S4 – S6).

#### *Model comparison*

To assess predictive performance of the different models we used a method which uses Pareto-smoothed Importance sampling to approximate the leave-one-out cross-validation estimate of the expected log predictive density for an out-of-sample data point (elpd), as used from the *loo* stan package[2]. We then compared the elpd from both models (constant vs time-varying) to obtain the $\mathrm{elpd}_{\mathrm{diff}}$. To determine whether this difference is significant we calculated the *z* score and compared it with the corresponding value from a standard normal distribution as recommended by Lambert [3].

$$Pr =\left( z\geq\frac{\mathrm{elpd}_{\mathrm{diff}}}{\mathrm{se}} \right)$$

### **Supplementary Figures**

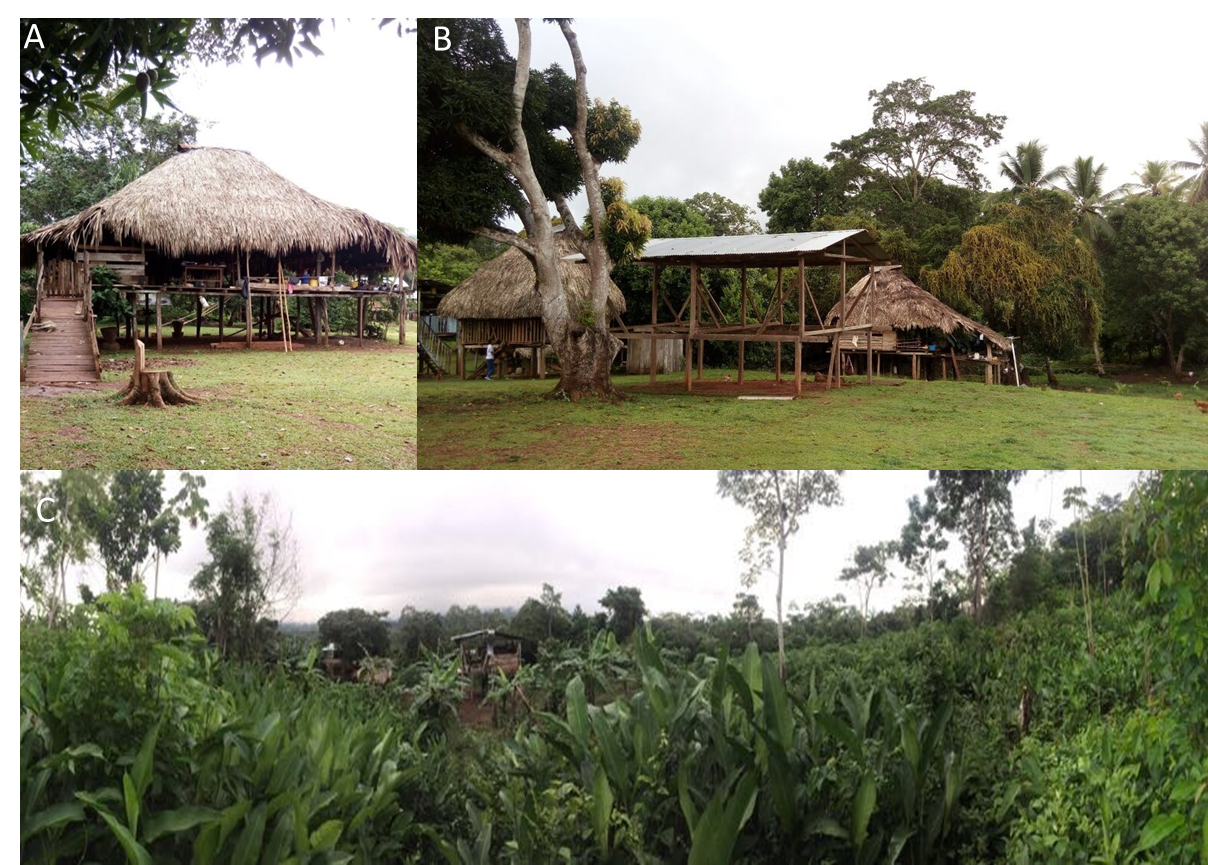

#### Figure S1. Photographs of the study sites

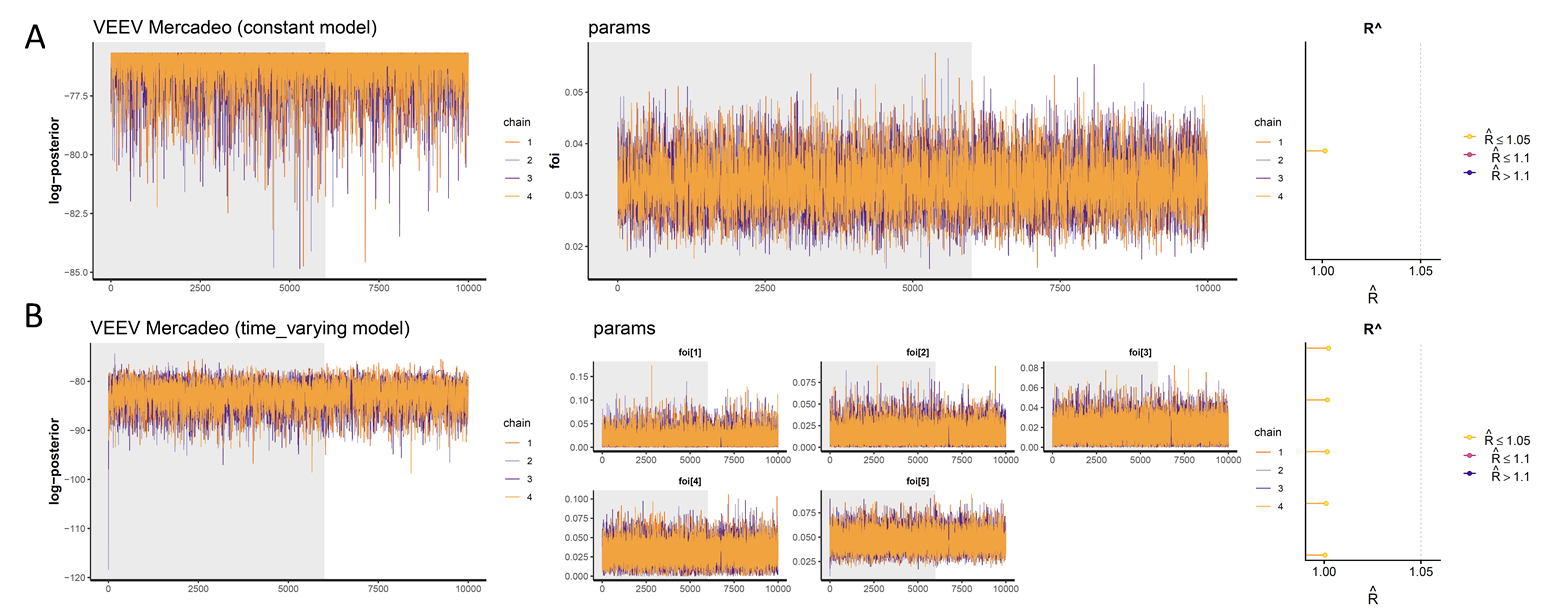

#### Figure S2. Convergence plots for VEEV FOI models in Mercadeo

Convergence plots for the FOI models for VEEV in Mercadeo, presenting the log posterior, posterior distribution of the parameters and $Rˆ$ values, for A) the constant FOI model and B) the time-varying FOI model

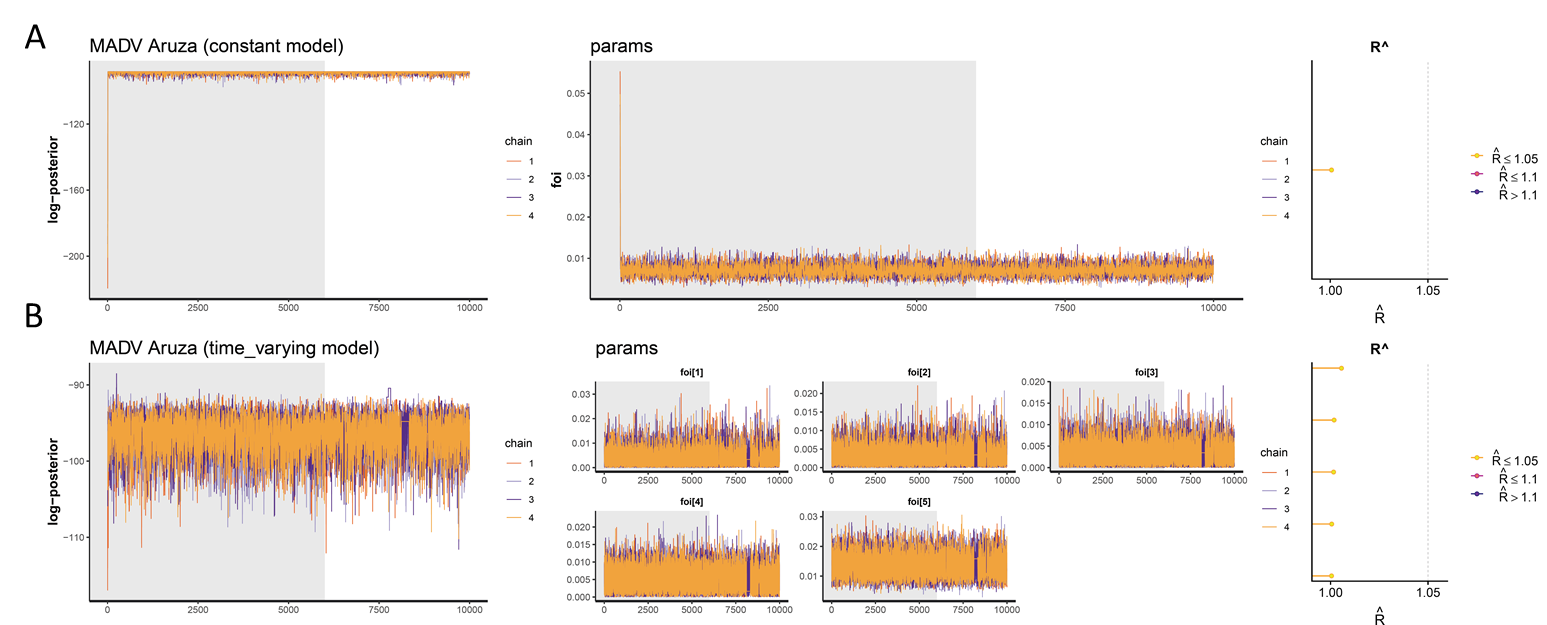

#### Figure S3. Convergence plots for MADV FOI models in Aruza

Convergence plots for the FOI models for MAVD in Aruza, presenting the log posterior, posterior distribution of the parameters and $Rˆ$ values, for A) the constant FOI model and B) the time-varying FOI model

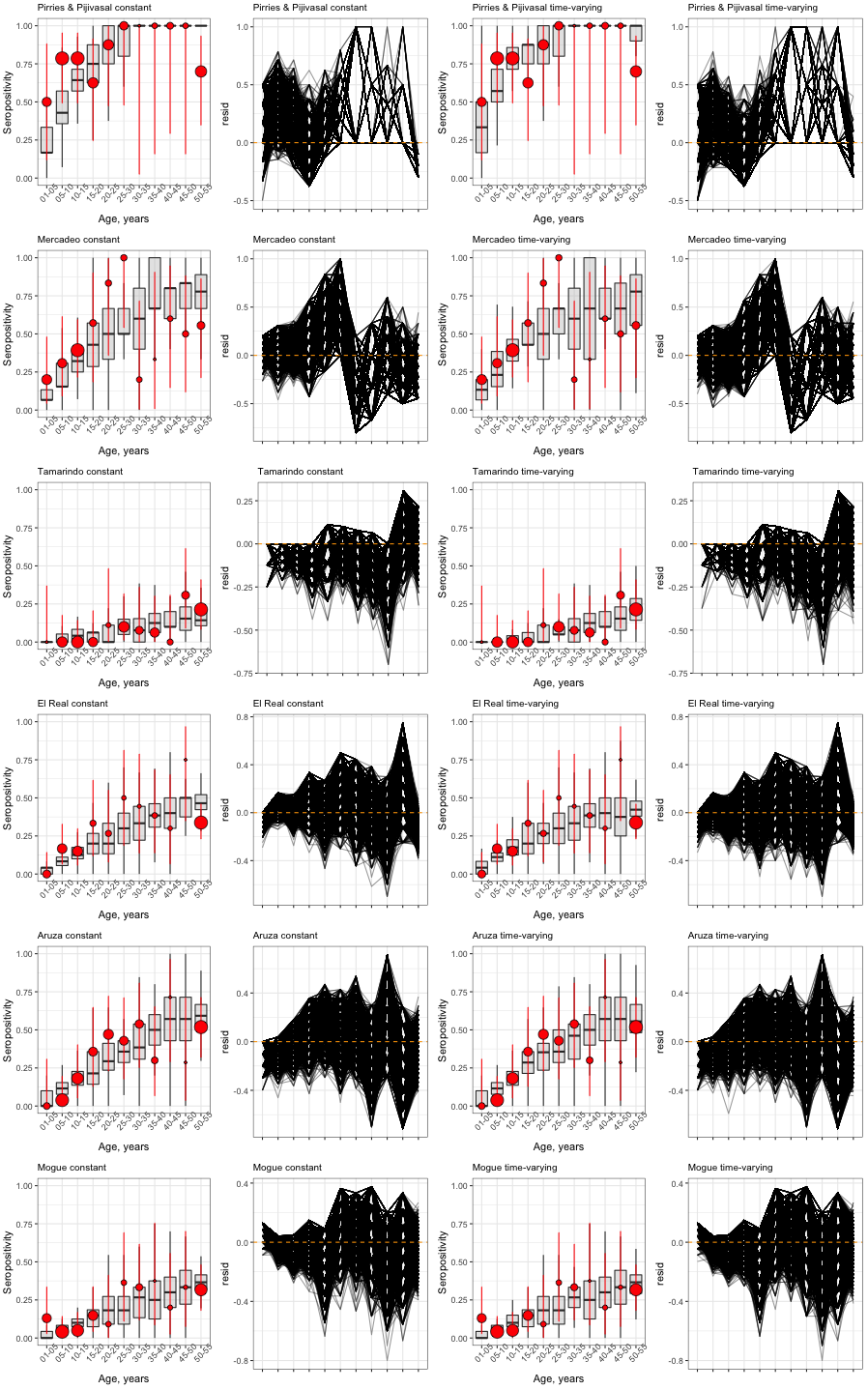

#### Figure S4. Posterior predictive checks and model residuals for VEEV for constant FOI models.

The left panels show the observed sero-prevalence data (red points represent mean and lines its 95% confidence intervals) and the model fitting (grey boxplots) The right panels show residuals of the model. Each row represents a location.

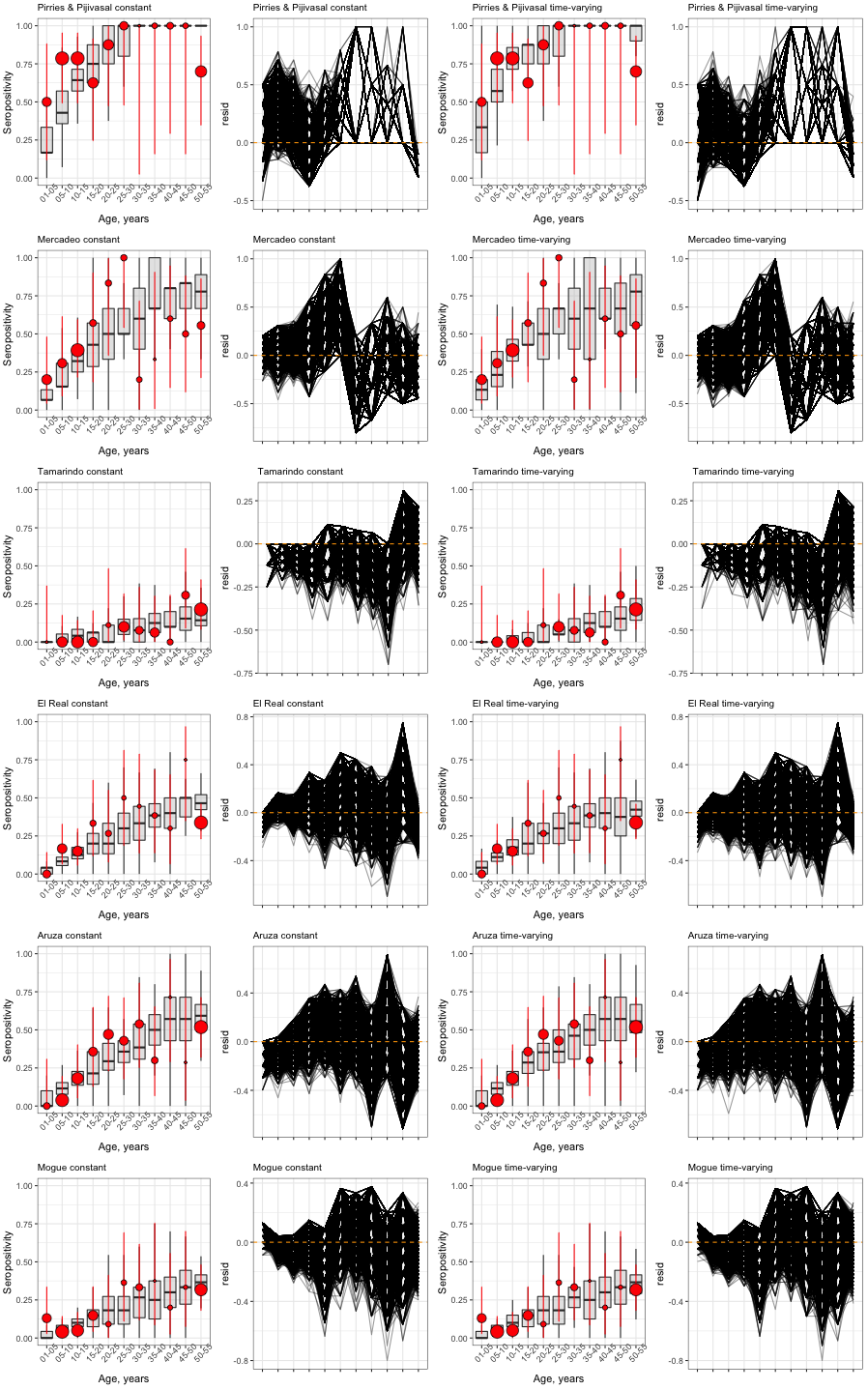

#### Figure S5. Posterior predictive checks and model residuals for VEEV for time-varying FOI models.

The left panels show the observed sero-prevalence data (red points represent mean and lines its 95% confidence intervals) and the model fitting (grey boxplots) The right panels show residuals of the model. Each row represents a location.

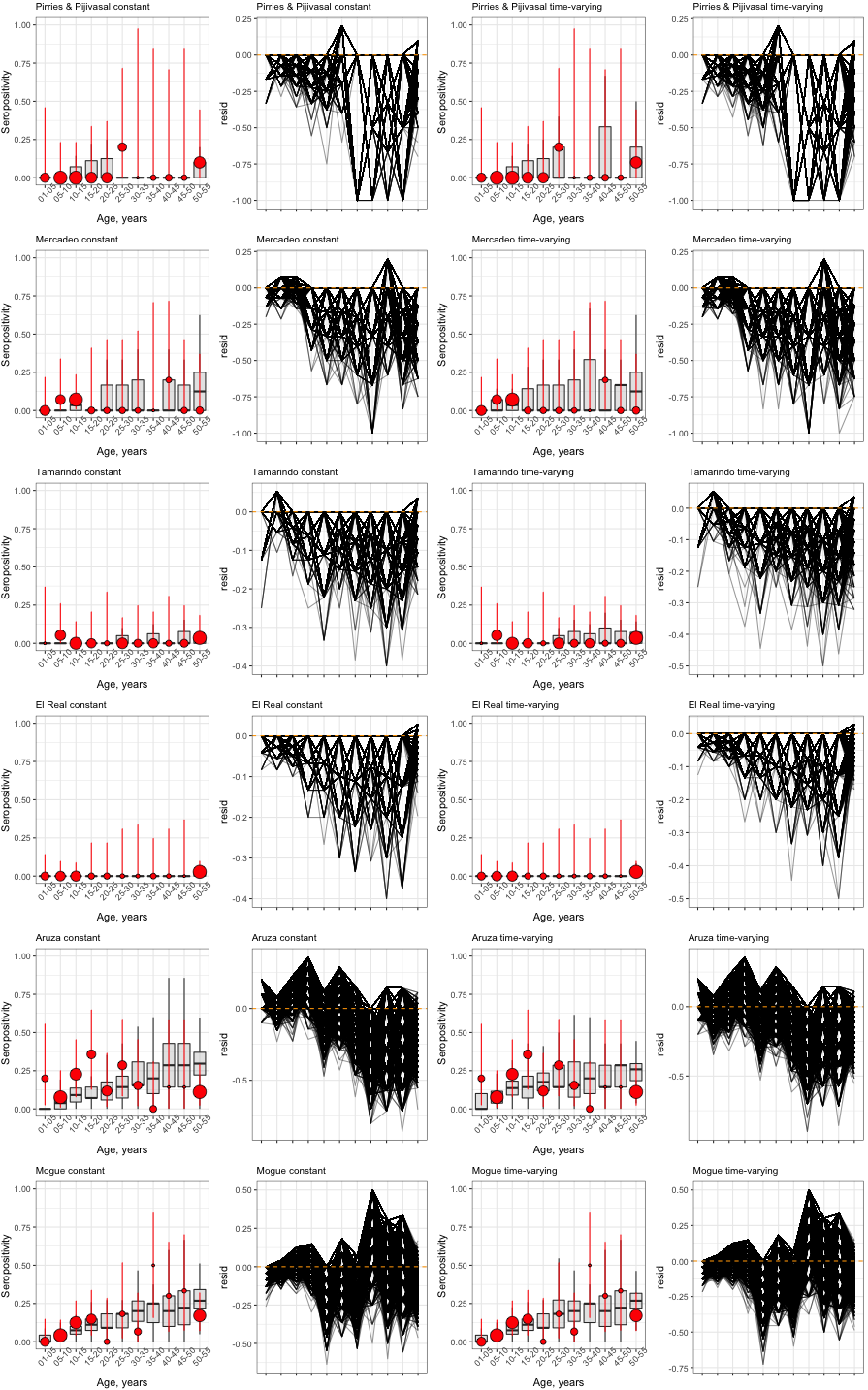

#### Figure S6. Posterior predictive checks and model residuals for MADV constant FOI models.

The left panels show the observed sero-prevalence data (red points represent mean and lines its 95% confidence intervals) and the model fitting (grey boxplots) The right panels show residuals of the model. Each row represents a location.

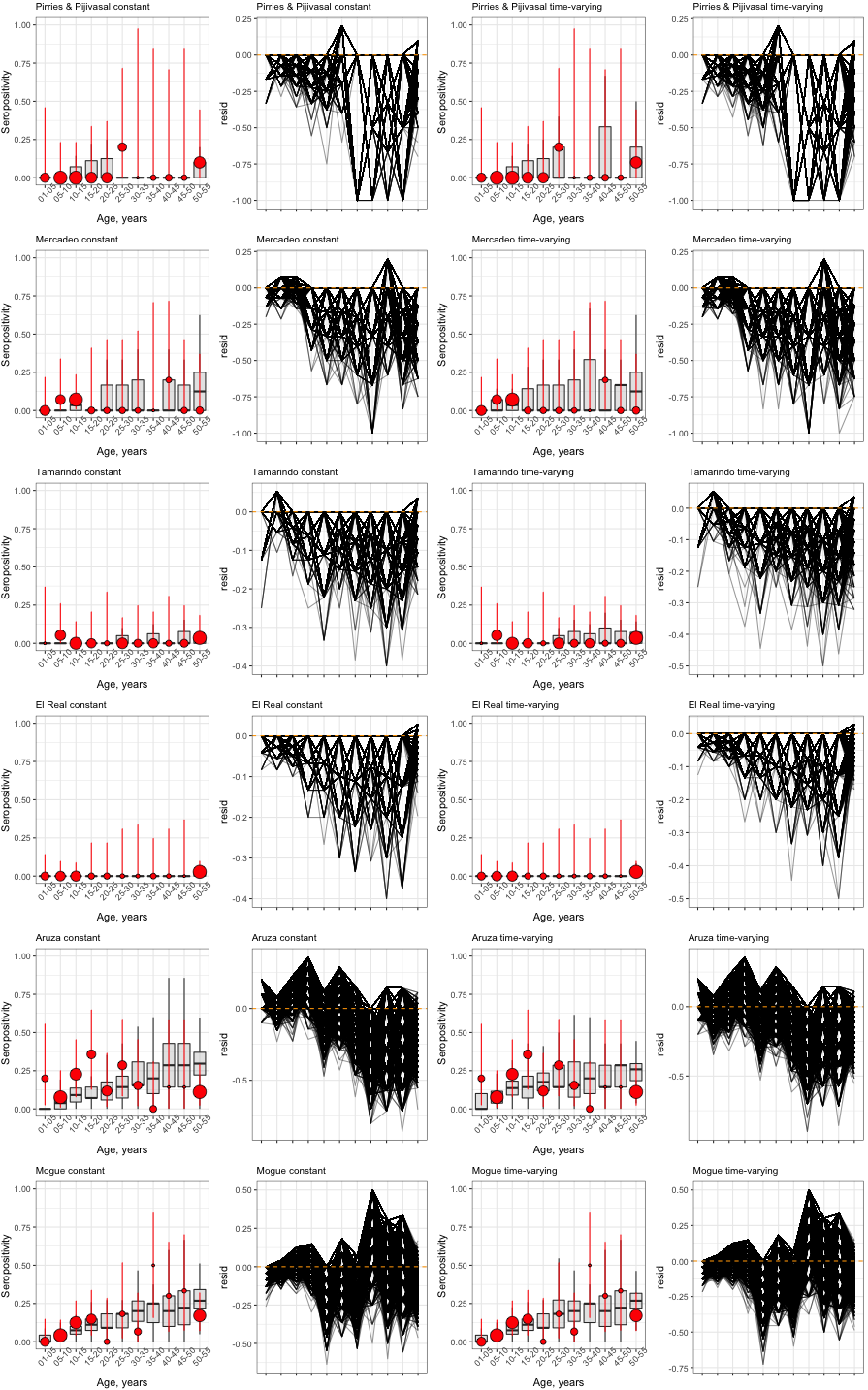

#### Figure S7. Posterior predictive checks and model residuals for MADV time-varying FOI models.

The left panels show the observed sero-prevalence data (red points represent mean and lines its 95% confidence intervals) and the model fitting (grey boxplots) The right panels show residuals of the model. Each row represents a location.

3.	Lambert B. A student’s guide to Bayesian statistics.
